# Protein biomarkers for response to XPO1 inhibition in hematologic malignancies

**DOI:** 10.1101/2022.12.15.22283531

**Authors:** Tulasigeri M. Totiger, Sana Chaudhry, Elgilda Musi, Jumana Afaghani, Skye Montoya, Frank Owusu-Ansah, Stanley Lee, Gary Schwartz, Virginia Klimek, Justin Taylor

## Abstract

XPO1 (Exportin-1) is the nuclear export protein responsible for the normal shuttling of several proteins and RNA species between the nucleocytoplasmic compartment of eukaryotic cells. XPO1 recognizes the nuclear export signal (NES) of its cargo proteins to facilitate its export. Alterations of nuclear export have been shown to play a role in oncogenesis in several types of solid tumor and hematologic cancers. Over more than a decade, there has been substantial progress in targeting nuclear export in cancer using selective XPO1 inhibitors. This has resulted in recent approval for the first-in-class drug selinexor for use in relapsed, refractory multiple myeloma and diffuse large B-cell lymphoma (DLBCL). Despite these successes not all patients respond effectively to XPO1 inhibition and there has been lack of biomarkers for response to XPO1 inhibitors in the clinic. Using hematologic malignancy cell lines and samples from patients with myelodysplastic neoplasms treated with selinexor, we have identified XPO1, NF-κB(p65), MCL-1 and p53 protein levels as protein markers of response to XPO1 inhibitor therapy. These markers could lead to the identification of response upon XPO1 inhibition for more accurate decision making in the personalized treatment of cancer patients undergoing treatment with selinexor.

## Introduction

XPO1 is a nuclear exporter responsible for exporting proteins that contain a nuclear export signal (NES) out of the nucleus to the cytoplasm^1-7^. XPO1 contributes to normal homeostasis of eukaryotic cells by regulating the export of key proteins,^5^ but alterations of *XPO1* promote oncogenesis and are associated with decreased survival in cancer^8-11^. Selinexor is a selective inhibitor of XPO1 that was recently FDA-approved for multiple myeloma and diffuse large B-cell lymphoma. However, not all patients respond effectively to XPO1 inhibition, and there is a lack of biomarkers of response to XPO1 inhibitors in advanced-phase clinical trials^12^. Here, we have identified XPO1, MCL-1, NF-κB and p53 expression as potential predictive biomarkers of response to XPO1 inhibitor therapy.

## Materials and Methods

### EC50 and synergy assays

Half-maximal effective concentration (EC50) assays were performed for selinexor or venetoclax with dimethyl sulfoxide (DMSO) as vehicle. Lymphoma cells (SUDHL-6, SUDHL-16, FARAGE, SUPHD1, L428) were treated in triplicate at 10,000 cells per well in 96-well plates (Costar) for 72 hours. Cell viability was measured by CellTiter-Glo (Promega). Synergy assays were performed for 72 hours utilizing 6×6 combination matrix in 96-well plates (Costar). Bliss synergy scores were calculated using SynergyFinder software.

### Drug treatment and immunoblots

Nalm6 wild type (WT) and Nalm6 *XPO1* E571K cells were seeded in 6-well plates and treated with 200nM of selinexor or 1μM of venetoclax in monotherapies or in combination for 24hrs. Cell lysates were collected using IP lysis buffer (Thermo Fisher, Cat# 87788) containing 0.1% EDTA and phosphatase inhibitor. 10 μg of protein was loaded on a 4-20% gel (Bio-Rad) and transferred onto a PVDF membrane. The antibodies used were XPO1(H-300) (Santa-Cruz, Cat# 55955), p53(DO-1, Santa Cruz, Cat# 126), NF-κB (Cell Signaling, Cat# D14E12), Anti-Rabbit MCL-1(Cell Signaling, Cat# D5V5L) and Beta-Actin (Sigma, Cat# A1978). Signal intensity quantification was performed by using ImageJ software.

### Ex vivo studies in patient samples

Patients with myelodysplastic syndromes (MDS, n=19) and oligoblastic acute myeloid leukemia (AML with 20-30% bone marrow blasts; n=4) who were refractory to hypomethylating agents were treated with selinexor monotherapy on a single center (MSKCC) IRB-approved clinical study as previously described^11^. Responses were assessed as per the modified International Working Group MDS response criteria^13^. Immunoblots were run as described above using p53 (Santa Cruz, Cat# 263), p21 (Cell Signaling, Cat# S947), MCL-1 (Cell Signaling, Cat# 4572), XPO1 (Santa Cruz, Cat# 5595 (H-300)], NF-κB (Cell Signaling, Cat# 4764) and GAPDH (Cell Signaling, Cat #2118) antibodies.

## Results

### XPO1 inhibition effectively killed hematologic malignant cell lines and cooperated with BCL-2 inhibition in combination

EC50 values of selinexor or venetoclax monotherapies in lymphoma cell lines (**Figure 1A)** revealed that *XPO1* mutant cells SUDHL-16 *XPO1* E571K were significantly more sensitive to selinexor (EC50 = 24nM) when compared to *XPO1* wild type (WT) cell lines SUDHL-6 and Farage (EC50 = 144nM and 41nM, respectively). SUDHL-16 *XPO1* E571K and SUPHD1 *XPO1* E571K are also sensitive to venetoclax (12nM and 10nM, respectively) when compared to SUDHL-6, Farage and L428 (EC50 = 26 nM, 47 nM and 287nM, respectively). Synergy assays **(Figure 1B, C)** demonstrated scores of 12.48 for Farage *XPO1 WT*,10.9 for SUDHL-6 *XPO1 WT* and 8.42 for SUPHD-1 *XPO1* E571K. These results provided evidence that XPO1 inhibition cooperates with BCL-2 inhibition in lymphoma cell lines and prompted us to look for the mechanisms underlying this synergy. Selinexor has a p53-dependent action^8^, while venetoclax has a non-p53 dependent action^14^, and these different mechanisms of action could explain an additive or synergistic effect. To date, there are no studies assessing the effect of venetoclax treatment on XPO1 cargoes. To address this, we evaluated the change in protein expression following treatment with selinexor and venetoclax as monotherapies and in combination.

**Figure 1.**
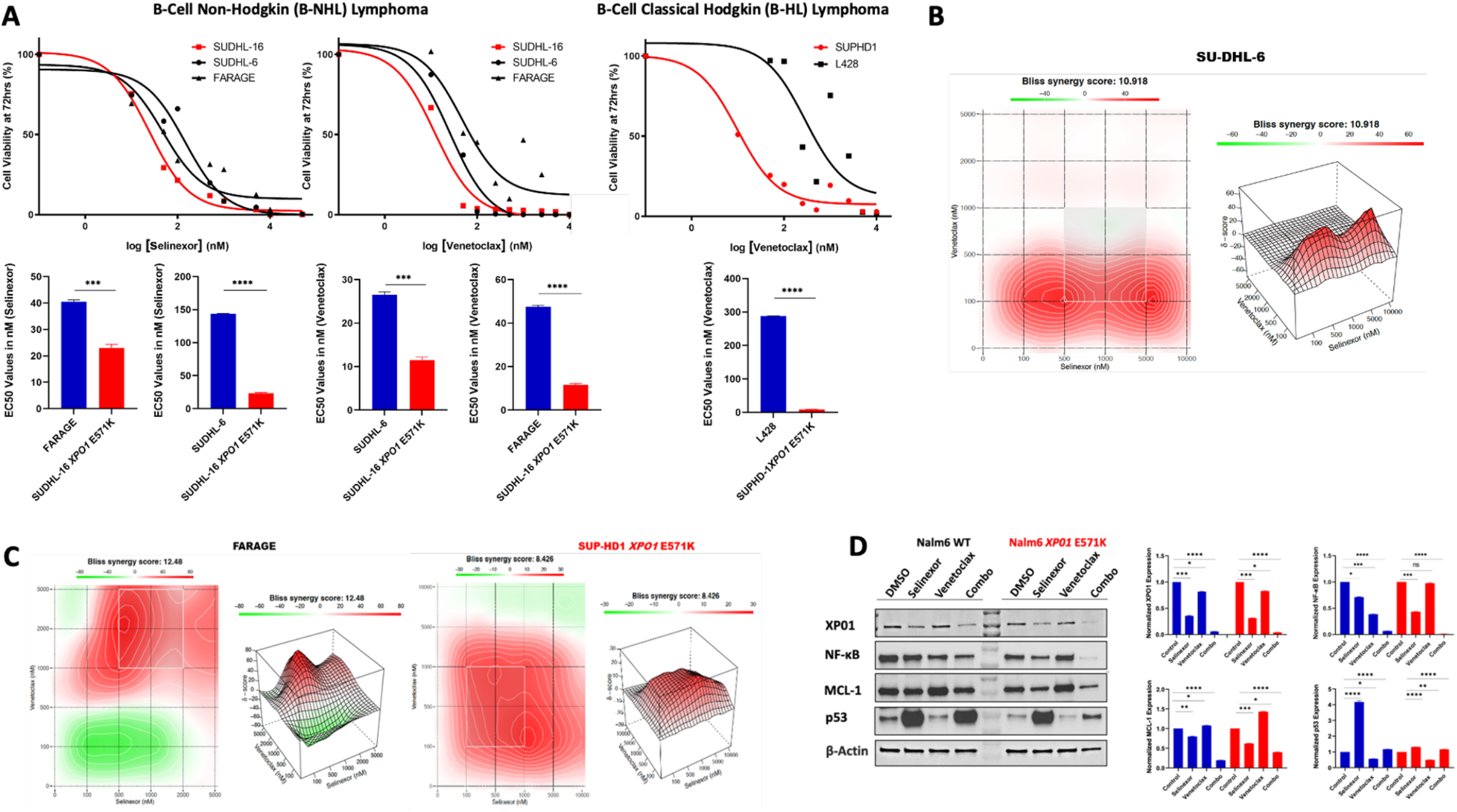
*In vitro* study showing response to selinexor, venetoclax and their combination. **A**. Cell viability curves (EC50) demonstrating the efficacy of selinexor and venetoclax against the indicated cell lines (left). Statistical analysis showing the significance of sensitivity of *XPO1* mutant cells against the *XPO1* wild type cells (right). **B**. Heat map panels showing the combination of selinexor and venetoclax in *XPO1* WT diffuse large B-cell lymphoma. **C**. Heat map for *XPO1 WT* and *XPO1* E571K Hodgkin’s lymphoma. **D**. Western blot demonstrating the effect of selinexor and venetoclax in monotherapies and in combination in B-cell leukemia WT and XPO1 mutant cell lines (left) and their densitometric analysis (right). Tukey’s multiple comparison test was used when comparing more than two groups and T-test was used for two group comparisons (****P< 0.0001, ***P<0.001, **P<0.01, *P<0.05).

The western blot results revealed that XPO1 protein, as well as NF-κB, MCL-1 and p53 protein levels, were affected by selinexor and venetoclax both as monotherapies and in combination **(Figure 1D)**. In particular, XPO1 protein levels were decreased in the *XPO1* mutant cell line (Nalm6 *XPO1* E571K) and its wild type counterpart (Nalm6 *XPO1* WT) following selinexor treatment and this suppression was further enhanced with the addition of venetoclax. With venetoclax monotherapy there was an induction of MCL-1 protein expression in both lines, whereas selinexor suppressed expression. Furthermore, the combination therapy was able to prevent induction of MCL-1 in both cell lines, essentially back to levels seen with selinexor alone. The response of NF-κB protein expression was decreased with selinexor treatment and with combination therapy particularly in the mutant cell line. Finally, p53 induction was quite dramatic with selinexor in both cell lines and there was only slight reduction with the combination therapy.

### MDS patients are sensitive to *XPO1* inhibition and exhibited similar pattern of protein profile response as seen *in vitro*

In this study, samples required for protein analysis were available from 13 patients with MDS. In these patients, XPO1 protein inhibition by selinexor showed differential responses across response categories, including marrow samples from complete remission (mCR; n=6), progressive disease (PD; n=4) and stable disease (SD; n=3) (**Figure 2**). For most responders **(Figure 2A)** XPO1 protein was either not present at baseline (Karyo-01 and Karyo-34), decreased over time with treatment (Karyo-31 and Karyo-12) or was found to be persistently low (Karyo-08). This contrasts with the progressors **(Figure 2B)** in which XPO1 protein expression in 3 of the 4 patients was inducible and remained elevated over time (Karyo-03, Karyo-09, and Karyo-22). Additionally, NFκB protein expression measured upon XPO1 inhibition, was found to generally follow the expression pattern of XPO1 (5 out of 6 in mCR, 3 out 4 in PD and 3 out of 3 in SD). This data is similar to the results seen *in vitro*, which is consistent with reports suggesting that inhibition of XPO1 modulates NF-kB signaling. There was no clear relationship between MCL-1 expression and clinical benefit. **(Figure 2A**; Karyo-12, Karyo-08 and Karyo-21) Lastly, there was no clear correlation between p53 expression and response. In fact, overall p53 expression was low to non-detectable in most patients (**Figure 2B**; Karyo-09, Karyo-03, and Karyo-22, **Figure 2C**; Karyo-19).

**Figure 2.**
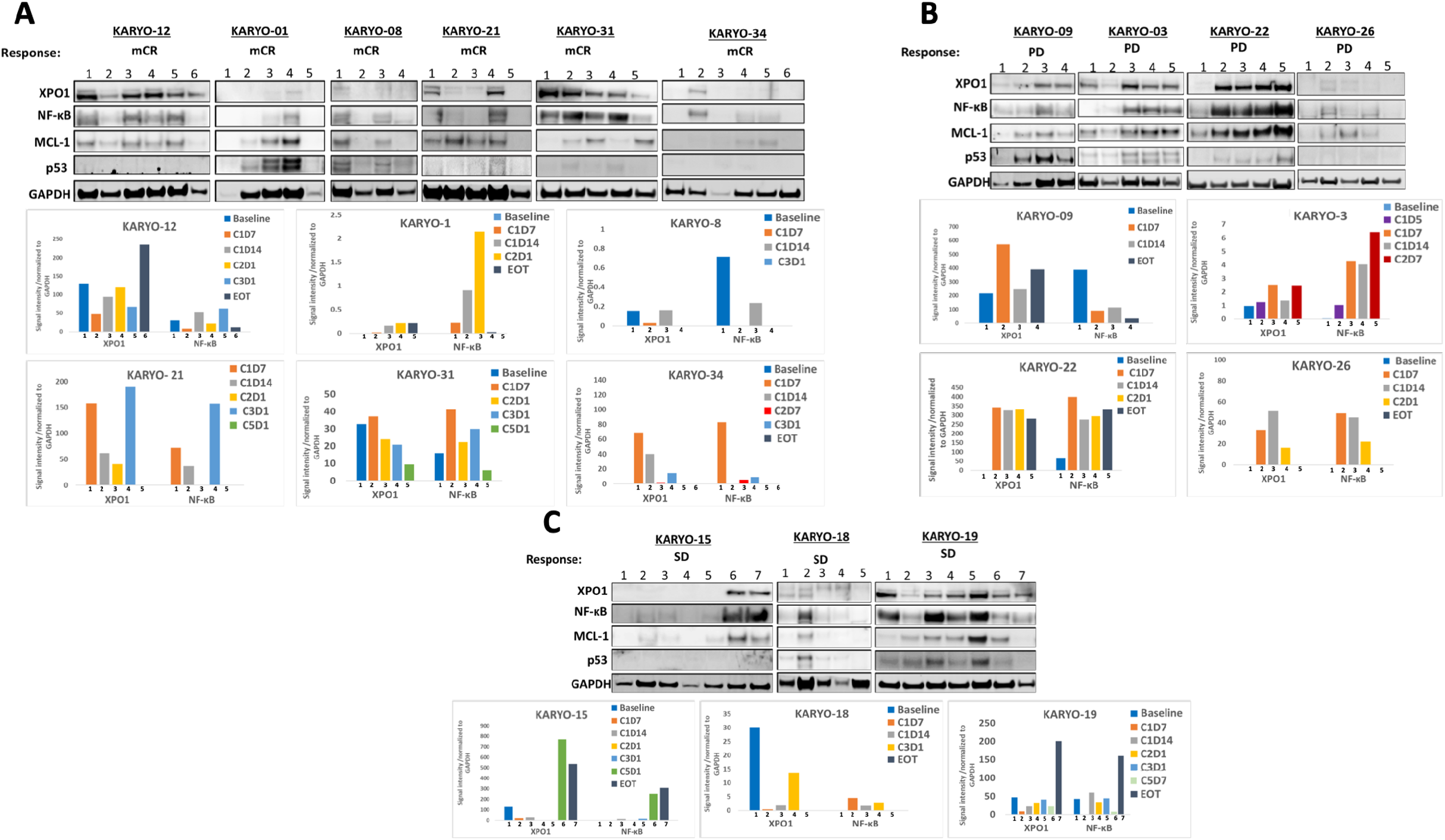
Western blot and densitometric analysis of serial samples from MDS patients treated with selinexor. Western blot analysis performed on serial samples from **A**. Patients who achieved marrow complete remission (mCR) **B**. Patients with progressive disease (PD) as best overall response **C**. Patients who achieved stable disease (SD). Densitometric analysis of the respective WB data from mCR, PD and SD patient samples are shown below the blots. Abbreviations: C=Cycle, D=Days, for example in C1D1, C1 represents cycle 1 and D1 represents day 1 of treatment. EOT=end of treatment.

## Discussion

Previous studies have shown that XPO1, MCL-1 and NF-κB protein levels are decreased upon XPO1 inhibition^14^ and XPO1 inhibition induces nuclear retention of p53 in solid tumors^15^. Hence, this prompted us to look at these potential predictive biomarkers of response to XPO1 inhibitor therapy *in vitro* and *in vivo*. Our *in vitro* studies confirmed that decreases in XPO1, MCL-1 and NF-κB protein levels and induction of p53 levels correlate with response to selinexor in monotherapy or in combination therapy. The clinical data with selinexor monotherapy showed differential responses of protein expression pattern on XPO1 inhibition across mCR, PD and SD patients with a trend in the mCR patients in favor of either suppression or lack of induction of XPO1 protein expression, especially at early time points when compared to the patients with PD. These results should be interpreted cautiously due to small sample size. However, these results represent the first attempt to correlate protein biomarkers with response to XPO1 inhibition in hematologic malignancies and should be further validated on a larger sample size. This could lead to the identification of easily measurable biomarkers of response for XPO1 inhibition that would allow more precise decision making for patients receiving this type of cancer therapy.

## Data Availability

All data will be made available upon reasonable request to the corresponding author.

## Author Contributions

TMT, SC, EM, JA, FOA, SL, SM and MA performed experiments and analysis. GS, VK, and JT conceptualized and supervised the work. TMT and JT wrote the original draft. All authors reviewed, edited and approved the final manuscript.

## Acknowledgements

JT is funded by the NCI/NIH (K08CA230319), the Doris Duke Charitable Foundation and the Edward P. Evans Foundation.

## Data Statement

All data will be made available upon reasonable request to the corresponding author.

## References

1. Stade K, Ford CS, Guthrie C, Weis K. Exportin 1 (Crm1p) is an essential nuclear export factor. Cell. 1997;90(6):1041–1050.

2. O’Reilly AJ, Dacks JB, Field MC. Evolution of the karyopherin-β family of nucleocytoplasmic transport factors; ancient origins and continued specialization. PLoS One. 2011;6(4):e19308.

3. Tran EJ, Bolger TA, Wente SR. SnapShot: nuclear transport. Cell. 2007;131(2):420.

4. Chook YM, Süel KE. Nuclear import by karyopherin-βs: recognition and inhibition. Biochim Biophys Acta. 2011;1813(9):1593–1606.

5. Castanotto D, Lingeman R, Riggs AD, Rossi JJ. CRM1 mediates nuclear-cytoplasmic shuttling of mature microRNAs. Proc Natl Acad Sci U S A. 2009;106(51):21655–21659.

6. Xu D, Grishin NV, Chook YM. NESdb: a database of NES-containing CRM1 cargoes. Mol Biol Cell. 2012;23(18):3673–3676.

7. Xu D, Farmer A, Collett G, Grishin NV, Chook YM. Sequence and structural analyses of nuclear export signals in the NESdb database. Mol Biol Cell. 2012;23(18):3677–3693.

8. Taylor J, Sendino M, Gorelick AN, et al. Altered Nuclear Export Signal Recognition as a Driver of Oncogenesis. Cancer Discov. 2019;9(10):1452–1467.

9. Kirli K, Karaca S, Dehne HJ, et al. A deep proteomics perspective on CRM1-mediated nuclear export and nucleocytoplasmic partitioning. Elife. 2015;4.

10. Hart T, Chandrashekhar M, Aregger M, et al. High-Resolution CRISPR Screens Reveal Fitness Genes and Genotype-Specific Cancer Liabilities. Cell. 2015;163(6):1515–1526.

11. Taylor J, Mi X, Penson AV, et al. Safety and activity of selinexor in patients with myelodysplastic syndromes or oligoblastic acute myeloid leukaemia refractory to hypomethylating agents: a single-centre, single-arm, phase 2 trial. Lancet Haematol. 2020;7(8):e566–e574.

12. Walker CJ, Oaks JJ, Santhanam R, et al. Preclinical and clinical efficacy of XPO1/CRM1 inhibition by the karyopherin inhibitor KPT-330 in Ph+ leukemias. Blood. 2013;122(17):3034–3044.

13. Cheson BD, Greenberg PL, Bennett JM, et al. Clinical application and proposal for modification of the International Working Group (IWG) response criteria in myelodysplasia. Blood. 2006;108(2):419–425.

14. Attiyeh EF, Maris JM, Lock R, et al. Pharmacodynamic and genomic markers associated with response to the XPO1/CRM1 inhibitor selinexor (KPT-330): A report from the pediatric preclinical testing program. Pediatr Blood Cancer. 2016;63(2):276–286.

15. Senapedis WT, Baloglu E, Landesman Y. Clinical translation of nuclear export inhibitors in cancer. Semin Cancer Biol. 2014;27:74–86.

